# Combined Use of Stress Echocardiography and Cardiopulmonary Exercise Testing to Assess Exercise Intolerance after Acute Myocardial Infarction

**DOI:** 10.1101/2020.08.23.20169821

**Authors:** Krzysztof Smarz, Tomasz Jaxa-Chamiec, Beata Zaborska, Maciej Tysarowski, Andrzej Budaj

## Abstract

**Background:** Exercise capacity (EC) after acute myocardial infarction (AMI) influences prognosis, but the causes of its reduction are complex and not sufficiently studied.

**Methods:** We prospectively enrolled consecutive patients who underwent percutaneous coronary intervention for their first AMI with left ventricular ejection fraction (LV EF) >40% at least 4 weeks after AMI. We performed combined stress echocardiography and cardiopulmonary exercise testing (CPET-SE) using a semi-supine cycle ergometer to determine predictors of EC (peak oxygen uptake [VO2]).

**Results:** Among 81 patients (70% male, mean age 58 ± 11 years), 40% had AMI with ST-segment elevation, and 60% non ST-segment elevation, LV EF was 57 ± 7%; wall motion score index, 1.18 (IQR 1.06 – 1.31); peak VO2, 19.5 ± 5.4 mL/kg/min. Multivariate analysis revealed that parameters at peak exercise: heart rate (β = 0.17, p < 0.001), stroke volume (β = 0.09, p < 0.001), and arteriovenous oxygen difference (A-VO2Diff, β = 93.51, p < 0.001) were independently positively correlated with peak VO2, with A-VO2Diff being its strongest contributor.

**Conclusions:** In patients treated for AMI with normal/mildly reduced LV EF, EC is associated with peak peripheral oxygen extraction as well as peak heart rate and peak stroke volume. CPET-SE is a useful tool to evaluate decreased fitness in this group.

## Introduction

Reduced exercise capacity (EC) after acute myocardial infarction (AMI) is common and renders poor prognosis.^1-5^ In a study of 2896 patients with newly diagnosed ischemic heart disease that included 1064 patients after AMI, EC assessed before cardiac rehabilitation was significantly decreased at roughly 60% of age-matched norms for healthy individuals without heart diseases.^3^ Contributors of reduced EC after MI are complex and can include cardiac ischemic injury, systolic and diastolic dysfunction, functional mitral regurgitation, chronotropic incompetence, but also peripheral muscle dysfunction.^6,7^ Deconditioning during the recovery period after AMI and can result in changes within the skeletal muscles similar to those observed in chronic heart failure.^8^ Resting left ventricular function parameters including LV EF have been shown to poorly corelate with EC, therefore other mechanisms such as peripheral factors or LV function during exercise needs to be investigated.^7,9-12^

Simultaneously performed stress echocardiography and cardiopulmonary exercise testing (CPET-SE) enables noninvasive assessment of cardiac function and peripheral oxygen extraction. It is an emerging diagnostic method with considerable potential in cardiology, especially in evaluation of the predictors of exercise intolerance.^13-18^ However, this strategy has been predominantly used in studies of patients with heart failure.^13,14,17,18^ Parameters that play crucial role in EC in patients after AMI, have not been sufficiently studied. The present study aims to assess the determinants of EC using CPET-SE in patients treated for AMI with normal or mildly reduced LV EF at least 4 weeks after AMI.

## Methods

### Study population

We prospectively enrolled all consecutive patients aged over 18 years who underwent percutaneous coronary intervention for their first AMI between October 2015 and January 2019 at the Department of Cardiology, Centre of Postgraduate Medical Education, Grochowski Hospital in Warsaw, Poland.

Flow chart of the study is presented in Figure 1. Study exclusion criteria were: previous AMI, history or presence of symptomatic congestive heart failure, permanent atrial fibrillation or atrial flutter, pulmonary disease with decreased forced expiratory volume in 1st second (FEV1) or vital capacity (VC), heart surgery, peripheral nerve and musculoskeletal disorder, peripheral vascular disease with intermittent claudication, stroke with residual deficits, left ventricular ejection fraction <40% at least 4 weeks after AMI, residual coronary artery stenosis (> 50%) after percutaneous coronary intervention, anemia (hemoglobin < 12 g/dL), decompensated thyroid disease, chronic kidney disease (creatinine clearance <30 mL/min), hemodynamically significant valvular defects, pulmonary hypertension, hypertrophic cardiomyopathy with left ventricular outflow tract obstruction, poor echocardiographic acoustic window and lack of informed consent, exercise-induced ischemia, pulmonary limitations of exercise, respiratory exchange ratio (RER) at peak exercise <1.05.

**Figure 1.**
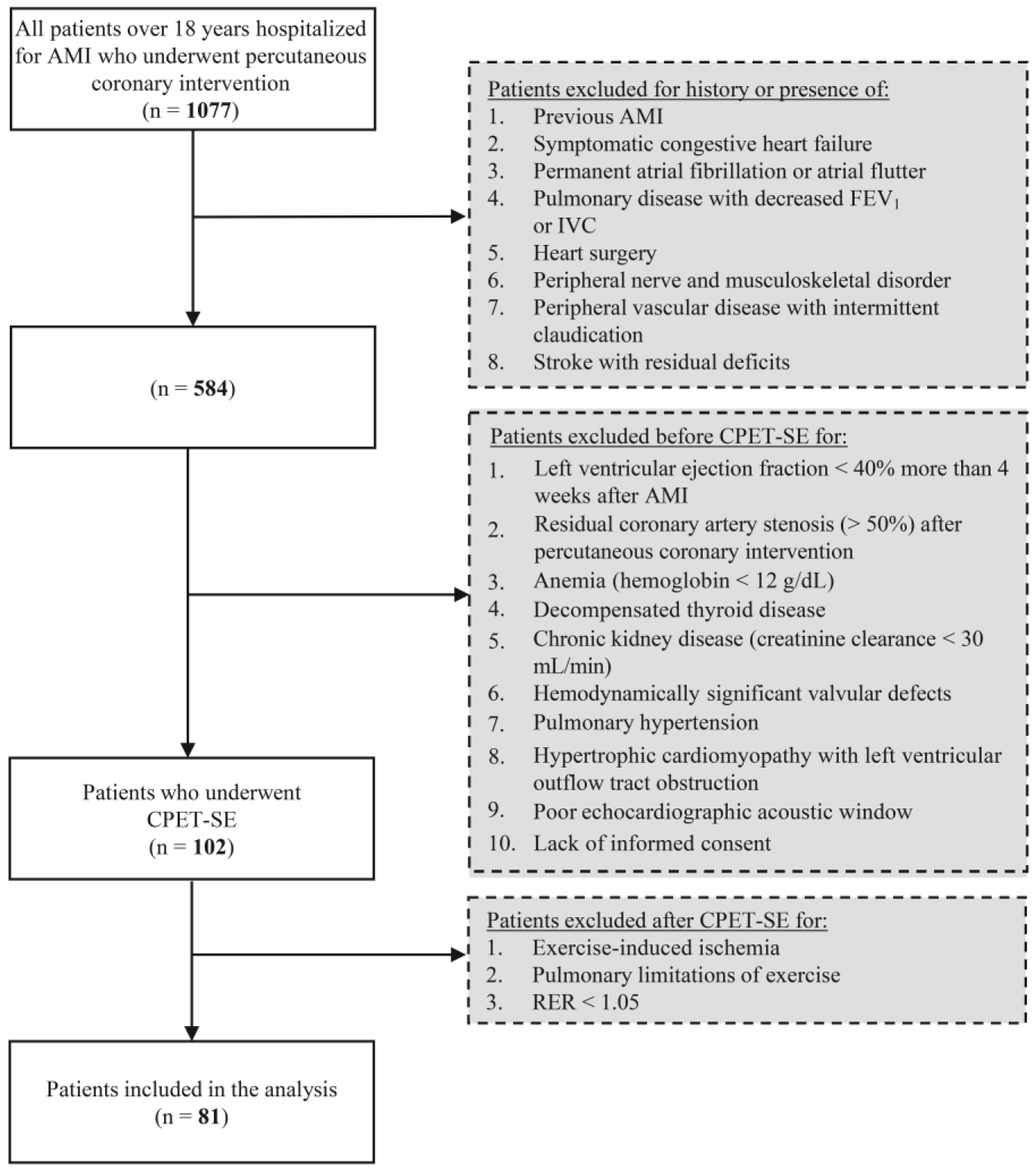
Study flow chart. Abbreviations: AMI, acute myocardial infarction; CPET-SE, combined stress echocardiography and cardiopulmonary exercise testing, FEV 1, forced expiratory volume in the first second; IVC, inspiratory vital capacity

We collected data on demographic characteristics, medical history and treatments as baseline characteristics. Self-assessed physical activity prior to AMI was categorized as low, moderate or high according to International Physical Activity Questionnaire.^19^

### Cardiopulmonary exercise testing

We performed symptom-limited cardiopulmonary exercise tests using a Schiller Cardiovit CS-200 (Schiller, Baar, Switzerland) and an Ergo Spiro adapter (Ganshorn, Niederlauer, Germany) with patients on a semi-supine cycle ergometer eBike EL (Ergoline GmbH, Bitz, Germany). Volumetric and gas calibration was performed daily before the tests. Volumetric calibration with respect to current temperature, relative air humidity and atmospheric pressure was carried out using a standard 2-L syringe. Gas calibration was performed using a standard gas mixture containing 15% oxygen, 6% carbon dioxide and 79% nitrogen. In all cases, we used a ramp protocol with an incremental load of 12.5 watts/min. All patients were familiar with the exercise protocol and were encouraged to exercise at maximal effort (≥8 points using the 10-point Borg scale).^20^ All exercise tests were supervised and analyzed according to the guidelines of the American College of Cardiology/American Heart Association and the American Thoracic Society/American College of Chest Physicians.^21^"^25^ During the stress test, we assessed the clinical and hemodynamic status of the patient and recorded electrocardiograms (12 leads) and ventilation and gas exchange parameters. The peak oxygen uptake (VO2), averaged from the last 20 s of exercise in mL/kg/min. The anaerobic threshold was calculated using a dual method approach (V-slope and ventilatory equivalent methods). Maximum predicted oxygen uptake was calculated according to the Wasserman/Hansen equations.^26^

The systolic and diastolic blood pressure and heart rate were recorded at rest and at peak exercise during the test, and the chronotropic index and percent of maximum predicted heart rate at peak exercise was calculated.^24^ The maximum predicted heart rate was calculated as 220 – age.^27^ Chronotropic incompetence was recognized as chronotropic index <80% for patients not on beta-blockers and ≤62% for patients on beta-blockers.^24^ Recorded electrocardiographic parameters included the presence or absence of ischemic changes, arrhythmias and conduction abnormalities according to the American Heart Association.^24^

Other analyzed cardiopulmonary exercise testing parameters included ventilatory efficiency (VE/VCO2 slope) and breathing reserve (BR) at peak exercise calculated as the percentage of maximum voluntary ventilation: [(maximum voluntary ventilation – minute ventilation at peak exercise)/maximum voluntary ventilation] x 100. Resting spirometry was used to evaluate FEV 1 and the inspiratory VC.

### Stress echocardiography

Echocardiography was performed at rest and at peak exercise using a VIVID 9 (General Electrics Medical System, Horten, Norway). Exercise echocardiography was carried out simultaneously with the cardiopulmonary exercise testing. Resting echocardiograms were recorded before the start of the exercise test. Echocardiographic images were recorded at peak exercise, immediately after reaching a respiratory exchange ratio of 1.00. Two-dimensional images were recorded in standard views. Left ventricular volumes were measured in 4 - and 2-chamber apical views and LV EF was calculated using the modified Simpson’s rule.^28^ Left ventricular systolic (s’) and early diastolic (e’) myocardial velocities were evaluated using pulsed-tissue Doppler at the basal segments of the interventricular septum and lateral wall and were presented as averaged values. Regional wall motion was assessed and graded using a 4-point scale where 1 represented normal and 4 represented dyskinetic with a 16-segment model and expressed as wall motion score index (WMSI). Mitral flow was assessed using pulse-tissue Doppler sample volume between mitral leaflets tips; early mitral inflow velocity (E), late (atrial) inflow velocity (A) and deceleration time.^29,30^ Stroke volume was calculated by multiplying the area of the left ventricular outflow tract at rest by the left ventricular outflow tract velocity-time integral (measured using pulsed-wave Doppler at rest and at peak exercise). Right ventricular systolic function was assessed by evaluating tricuspid annular plane systolic excursion (TAPSE) and right ventricular systolic myocardial velocity (RV s’) in 2-chamber apical view. The arteriovenous oxygen difference (A-VO2Diff) was calculated using the Fick equation as follows: VO2/cardiac output calculated from echocardiography.^13,14^

Measurements and recordings of echocardiographic parameters were carried out according to American Society of Echocardiography and European Association of Echocardiography recommendations.^28,29,31,32^ Images were analyzed off-line using EchoPAC PC software v. 110.0.x.

All CPET-SE examinations were performed and interpreted by one cardiologist experienced in stress echocardiography and cardiopulmonary exercise testing.

### Statistical methods

Data are presented as mean ± standard deviation or median (25th - 75th percentiles) for continuous variables. Categorical variables are presented as a number (percentage). Normality for all continuous variables was tested using the Shapiro–Wilk test. Group comparisons between continuous variables were performed using the Welch's t-test or Mann–Whitney U test, and the Fisher exact test or χ^2^ (chi-squared) test for categorical variables. We evaluated the determinants of EC using multivariate linear regression with peak VO2 as the dependent variable. To identify predictors of EC we initially performed univariate analysis to assess the associations between all rest and stress echocardiographic and cardiopulmonary variables and peak VO2. We chose the variables with univariate correlations with p value < 0.05. To reduce collinearity, we used correlation factor analysis to determine strongly correlated pairs of predictor variables (correlation coefficients ≥ 0.7). From those highly correlated pairs we included predictors with lower univariate p value (calculated in the first step, with peak VO2 as the dependent variable) into the analysis. We obtained 24 variables that we further introduced into stepwise regression model using Akaike Information Criterion to obtain final predictors of peak VO2 in the multivariate linear regression model. Variables with well-known effect on EC were forced into the model. Predictors are presented with ß-coefficient and 95% confidence intervals. All statistical tests were two-sided.

Statistical significance was accepted at α = 0.05. All statistical analyses were performed using R Statistical Software version 3.6.1.

### Ethical statement

This study was conducted in conformance with the requirements set out in the Declaration of Helsinki. All patients provided written consent to participate. The study and all its protocols were approved by the Bioethical Committee of the Centre of Postgraduate Medical Education.

## Results

Out of 102 eligible patients, after CPET-SE 81 patients (57 male, mean age 58 ± 11 years) were finally enrolled into analysis. Flowchart of the study is presented in Figure 1. Among these, 40% had AMI with ST segment elevation and 60% non-ST segment elevation. Almost 31% of the study population had diabetes mellitus or impaired glucose tolerance, 67% hypertension and 47% were smokers. All patients were on optimal medical therapy for AMI with 70 patients on beta-blockers. Beta-blockers were not withheld before the exercise tests. Clinical characteristics of the study population are presented in Table 1.

**Table 1.** Demographic and clinical characteristics during hospitalization for acute myocardial infarction.

|  | All patients<br>(n=81) |
| --- | --- |
| **Demographics** |  |
| Male sex, n (%) | 57 (70) |
| Age, years | 57.7 ± 11.0 |
| Body mass index, kg/m<sup>2</sup> | 27.5 ± 4.2 |
| **Comorbidity, n (%)** |  |
| Current smoking | 38 (47) |
| Past smoking | 11 (14) |
| Hypertension | 54 (67) |
| Hyperlipidemia | 63 (78) |
| Diabetes mellitus/Impaired glucose tolerance | 25 (31) |
| Paroxysmal atrial fibrillation | 2 (2.5) |
| Physical activity before myocardial infarction |  |
| Low | 17 (21) |
| Moderate | 41 (51) |
| High | 23 (28) |
| **Clinical characteristics** |  |
| STEMI, n (%) | 32 (39) |
| Inferior | 18 (22) |
| Lateral | 8 (10) |
| Posterior | 3 (4) |
| Anterior | 13 (16) |
| Right ventricular infarction, n (%) | 1 (1) |
| Non STEMI, n (%) | 49 (61) |
| Culprit lesion |  |
| Left main coronary artery | 0 (0) |
| Left anterior descending artery | 42 (52) |
| Circumflex artery | 19 (23) |
| Right coronary artery | 20 (25) |
| Troponin T maximal plasma concentration, ng/L | 784 (242 – 2216) |
| Hemoglobin at discharge, g/dL | 14.05 ± 1.21 |
| Creatinine at discharge, mg/dL | 0.85 ± 0.14 |
| Creatinine clearance - Cockcroft-Gault equation, mL/min | 108 ± 30 |
| Left ventricular ejection fraction, % | 53.7 ± 7.4 |
| **Medication at discharge, n (%)** |  |
| ACE-I/ARB | 78 (96) |
| Beta-blocker | 70 (86) |
| Aspirin | 80 (99) |
| Oral antiplatelet therapy | 79 (97) |
| Statin | 79 (97) |
| Nitrate | 4 (5) |
| Calcium channel blocker | 19 (23) |
| Diuretic | 22 (27) |

Note: Values represent mean ± SD, median (IQR) or number (%). Oral antiplatelet therapy: clopidogrel or ticagrelor.
Abbreviations: ACE-I, angiotensin converting enzyme inhibitors; ARB, angiotensin receptor blockers; CPET-SE, combined stress echocardiography and cardiopulmonary exercise testing; STEMI, acute myocardial infarction with ST segment elevation.

### Combined stress echocardiography and cardiopulmonary exercise testing

The mean time from AMI to CPET-SE was 60 ± 37 days. Mean peak VO2 was 19.5 ± 5.4 mL/kg/min (20.6 ± 4.8 mL/kg/min for men and 16.8 ± 5.8 for women). Most studied patients had reduced EC assessed as percent of predicted peak VO2 (89% had %VO2 peak <100%). The median RER at peak exercise was 1.14 (IQR 1.07 – 1.21). Mean BR at peak exercise was 53 ± 12% and no participants had a BR of <15% at peak exercise. Resting spirometry and exercise test parameters are presented in Table 2.

**Table 2.**
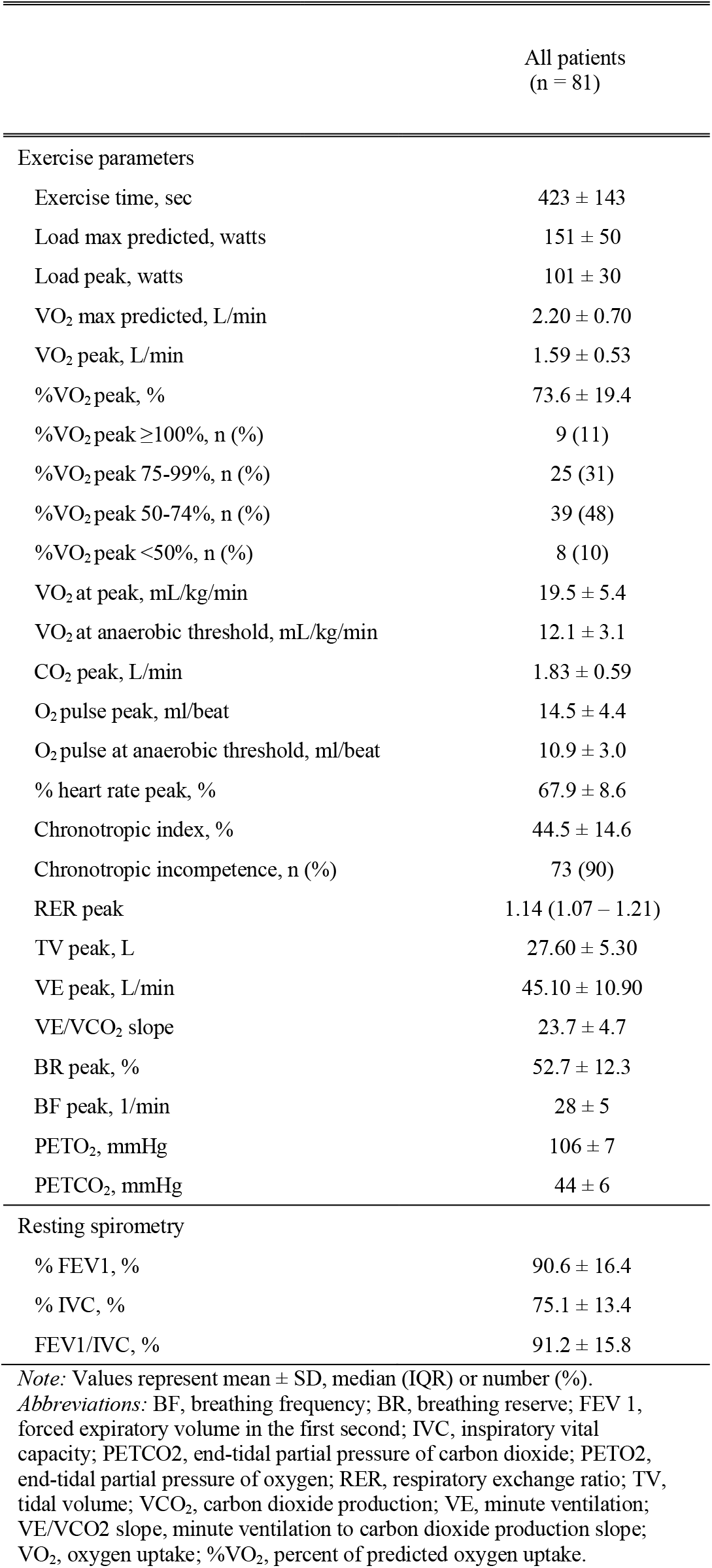
Cardiopulmonary parameters during combined exercise testing (CPET-SE)

Differences between parameters at rest and at peak exercise are presented in Table 4. As compared to rest, the mean heart rate and SBP increased and DBP decreased. Left ventricular stroke volume, LV EF, s’ increased and left ventricular end-systolic volume decreased with no changes in left ventricular end-diastolic volume. E/A mitral inflow velocity ratio, e’, E/e’ increased and deceleration time decreased during exercise. Right ventricular systolic function parameters: TAPSE, RV s’ and A-VO2Diff increased. There was no significant deterioration of mitral and tricuspid regurgitation during exercise.

### Determinants of exercise capacity

Univariate analysis (Table 5) revealed positive correlations between peak VO2 and male gender, heart rate peak, % heart rate, chronotropic index; FEV 1, e’ at rest and at peak, s’ at peak, and A-VO2Diff at rest, at peak and peak to rest difference. Age, LV EF at rest, DBP at peak and VE/VCO2 slope were negatively correlated with peak VO2.

Multivariate regression analysis revealed that only peak exercise parameters: heart rate, stroke volume, and A-VO2Diff were positively corelated with peak VO2. The A-VO2Diff at peak exercise was the strongest independent predictor of peak VO2 (partial R2 = 68%, Table 3). The univariate relationship between those variables and peak VO2 are presented in Figure 2.

**Table 3.** Multivariate analysis assessing predictors of exercise capacity measured as oxygen uptake at peak exercise (mL/min/kg).

| | $\beta$ - regression coefficient | 95% CI | p value |
| --- | --- | --- | --- |
| A-VO<sub>2</sub>Diff at peak, mL/dL | 93.5 | 77.9 - 109.1 | **<0.001** |
| Heart rate at peak, bpm | 0.17 | 0.12 - 0.22 | **<0.001** |
| Stroke volume at peak, mL | 0.09 | 0.05 - 0.14 | **<0.001** |
| SBP, mmHg | -0.03 | -0.06 - 0.00 | 0.058 |
| e' at rest, cm/s | 0.35 | -0.02 - 0.80 | 0.061 |
| LV s' at peak, cm/s | -0.01 | -0.21 - 0.90 | 0.219 |
| WMSI at rest | -2.6 | -6.81 - 1.61 | 0.222 |
| LV s' at rest, cm/s | -0.29 | -0.79 - 0.21 | 0.255 |
| Age, years | 0.03 | -0.04 - 0.11 | 0.358 |
| Stroke volume at rest, mL | -0.02 | -0.08 - 0.04 | 0.482 |
| e' at peak, cm/s | -0.16 | -0.50 - 0.19 | 0.373 |
| LV EF at peak, % | -0.01 | -0.08 - 0.07 | 0.895 |
| LV EF at rest, % | 0 | -0.10 - 0.10 | 0.983 |

Note: n = 81, R<sup>2</sup> = 81%, adjusted R<sup>2</sup> = 77%. Bold values indicate statistical significance.
Abbreviations: A-VO<sub>2</sub>Diff, arteriovenous oxygen difference; e', early diastolic myocardial velocity; LV EF, left ventricular ejection fraction; LV s', left ventricular systolic myocardial velocity; SBP, systolic blood pressure; WMSI, wall motion score index.

**Table 4.** Resting and at peak exercise parameters during combined stress echocardiography and cardiopulmonary exercise testing.

|  | Rest | Peak | p value |
| --- | --- | --- | --- |
| Heart rate, bpm | 67 ± 8 | 109 ± 14 | **<0.001** |
| SBP, mmHg | 127 ± 15 | 181 ± 22 | **<0.001** |
| DBP, mmHg | 74 ± 8 | 69 ± 12 | **0.002** |
| Stroke volume, mL | 79.0 ± 15.1 | 95.6 ± 19.9 | **<0.001** |
| WMSI | 1.18 (1.06 – 1.31) | 1.13 (1.06 – 1.26) | 0.211 |
| LV EF, % | 57.5 ± 7.6 | 66.3 ± 9.4 | **<0.001** |
| LVEDV, mL | 100.2 ± 30.0 | 97.3 ± 28.6 | 0.153 |
| LVESV, mL | 43.5 ± 17.7 | 34.0 ± 16.1 | **<0.001** |
| LV s', cm/s | 8.0 ± 1.7 | 10.2 ± 1.9 | **<0.001** |
| E/A mitral inflow velocity ratio | 1.02 ± 0.4 | 1.28 ± 0.45 | **<0.001** |
| Deceleration time, ms | 230 (200 – 280) | 172 (153 – 206) | **<0.001** |
| e', cm/s | 8.8 ± 2.2 | 12.2 ± 2.7 | **<0.001** |
| E/e' | 7 ± 2 | 8 ± 2 | **0.025** |
| TAPSE, cm | 2.24 ± 0.27 | 2.91 ± 0.51 | **<0.001** |
| RV s', cm/s | 12.2 ± 2.1 | 16.3 ± 2.8 | **<0.001** |
| Mitral regurgitation, n (%) |  |  |  |
| Mild | 48 (59.3) | 45 (55.6) | 0.227 |
| Moderate | 2 (2.5) | 7 (8.6) | 0.170 |
| Severe | 0 | 0 | 1 |
| Tricuspid regurgitation, n (%) |  |  |  |
| Mild | 30 (37.0) | 32 (39.5) | 0.746 |
| Moderate | 0 | 1 (1.2) | 1 |
| Severe | 0 | 0 | 1 |
| A-VO<sub>2</sub>Diff | 0.08 ± 0.03 | 0.15 ± 0.05 | **<0.001** |

Note: Values represent mean ± SD, median (IQR), or number (%). Bold values indicate statistical significance.
Abbreviations: A, late mitral inflow velocity; A-VO<sub>2</sub>Diff, arteriovenous oxygen difference; DBP, diastolic blood pressure; E, early mitral inflow velocity; e', early diastolic myocardial velocity; LV EF, Left ventricular ejection fraction; LV EDV, left ventricular end-diastolic volume; LV ESV, left ventricular end-systolic volume; LV s', left ventricular systolic myocardial velocity; RV s', right ventricular systolic myocardial velocity; SBP, systolic blood pressure; TAPSE, tricuspid annulus plane systolic excursion; WMSI, wall motion score index.

**Table 5.** Univariate analysis assessing predictors of exercise capacity measured as oxygen uptake at peak exercise (mL/min/kg).

| | R value | Regression Coefficient ± SE | p-value |
| --- | --- | --- | --- |
| Age, years | -0.16 | -0.16 ± 0.05 | **0.003** |
| Sex, male | | 3.84 ± 1.26 | **0.003** |
| Body mass index, kg/m<sup>2</sup> | -0.24 | -0.24 ± 0.14 | 0.097 |
| Creatinine clearance, mL/min | 0.18 | 0.04 ± 0.02 | 0.050 |
| Hemoglobin, g/dL | 0.12 | 0.80 ± 0.50 | 0.110 |
| Heart rate at rest, bpm | -0.03 | -0.01 ± 0.06 | 0.837 |
| Heart rate at peak, bpm | 0.42 | 0.19 ± 0.03 | **<0.001** |
| Heart rate at peak, percent predicted, % | 0.26 | 0.17 ± 0.06 | **0.002** |
| Chronotropic index, % | 0.36 | 0.15 ± 0.04 | **<0.001** |
| SBP at rest, mmHg | 0.06 | -0.04 ± 0.04 | 0.319 |
| DBP rest, mmHg | -0.04 | -0.02 ± 0.07 | 0.734 |
| SBP at peak, mmHg | 0.06 | 0.01 ± 0.02 | 0.785 |
| DBP at peak, mmHg | -0.19 | -0.11 ± 0.04 | **0.027** |
| VE/VCO<sub>2</sub> slope | -0.25 | -0.62 ± 0.11 | **<0.001** |
| FEV1, percent predicted, % | 0.40 | 3.64 ± 0.85 | **<0.001** |
| IVC, percent predicted, % | -0.28 | -4.63 ± 3.80 | 0.229 |
| Stroke volume at rest, mL | -0.01 | -0.00 ± 0.04 | 0.987 |
| Stroke volume at peak, mL | 0.09 | 0.03 ± 0.03 | 0.403 |
| WMSI at rest | -0.10 | -4.63 ± 3.80 | 0.229 |
| WMSI at peak | -0.12 | -6.23 ± 4.21 | 0.145 |
| LV EF at rest, % | -0.18 | -0.17 ± 0.07 | **0.037** |
| LV EF at peak, % | 0.03 | 0.01 ± 0.65 | 0.926 |
| LV EF peak – rest, % | 0.19 | 0.12 ± 0.06 | 0.075 |
| LV EDV at rest, mL | 0.21 | 0.03 ± 0.02 | 0.087 |
| LV EDV at peak, mL | 0.09 | 0.02 ± 0.02 | 0.437 |
| LV ESV at rest, mL | 0.23 | 0.06 ± 0.03 | 0.087 |
| LV ESV at peak, mL | 0.01 | 0.01 ± 0.03 | 0.844 |
| LV s' at rest, cm/s | 0.20 | 0.59 ± 0.35 | 0.098 |
| LV s' at peak, cm/s | 0.36 | 1.19 ± 0.31 | **<0.001** |
| E/A mitral inflow velocity ratio at rest | 0.12 | 0.91 ± 1.58 | 0.553 |
| E/A mitral inflow velocity ratio at peak | 0.16 | 1.79 ± 1.4 | 0.206 |
| e' at rest, cm/s | 0.22 | 0.57 ± 0.26 | **0.033** |
| e' at peak, cm/s | 0.25 | 0.49 ± 0.22 | **0.029** |
| E/e' at rest | -0.20 | -0.43 ± 0.28 | 0.118 |
| E/e' at peak | -0.02 | -0.09 ± 0.24 | 0.711 |
| TAPSE at rest, cm | 0.02 | 0.60 ± 2.23 | 0.789 |
| TAPSE at peak, cm | 0.15 | 2.35 ± 1.65 | 0.051 |
| TAPSE peak - rest, cm | 0.17 | 2.52 ± 1.24 | 0.052 |
| RV s' at rest, cm/s | -0.01 | 0.00 ± 0.27 | 0.991 |
| RV s' at peak, cm/s | 0.20 | 0.41 ± 0.22 | 0.065 |
| RV s' peak – rest, cm/s | 0.22 | 0.12 ± 0.12 | 0.312 |
| A-VO<sub>2</sub>Diff at rest, mL/dL | 0.37 | 62.10 ± 21.16 | **0.005** |
| A-VO<sub>2</sub>Diff at peak, mL/dL | 0.67 | 81.12 ± 9.98 | **<0.001** |
| A-VO<sub>2</sub>Diff peak - rest, mL/dL | 0.53 | 85.21 ± 13.25 | **<0.001** |

Abbreviations: A, late mitral inflow velocity; A-VO<sub>2</sub>Diff, arteriovenous oxygen difference; DBP, diastolic blood pressure; E, early mitral inflow velocity; e', early diastolic myocardial velocity; LV EF, Left ventricular ejection fraction; LV EDV, left ventricular end-diastolic volume; LV ESV, left ventricular end-systolic volume; LV s', left ventricular systolic myocardial velocity; RV s', right ventricular systolic myocardial velocity; SBP, systolic blood pressure; SE, standard error; TAPSE, tricuspid annulus plane systolic excursion; WMSI, wall motion score index; FEV1, forced expiratory volume in 1 second; IVC, inspiratory vital capacity.

## Discussion

Our study revealed that in post AMI patients, peripheral (non-cardiac mechanisms) may play significant role in decreasing EC. We found that in patients treated for AMI with normal or mildly reduced LV EF, peak A-VO2Diff is the strongest contributor to EC. Other parameters independently associated with EC were peak heart rate and peak stroke volume. Stroke volume and heart rate during exercise reflect cardiac function, while A-VO2Diff reflects peripheral oxygen extraction. Thus, stress echocardiographic parameters are complementary to results obtained from cardiopulmonary exercise testing. It also implies the importance of peripheral factors, such as oxygen consumption by working muscles, for the EC of patients after AMI with normal or mildly reduced LV EF.

**Figure 2.**
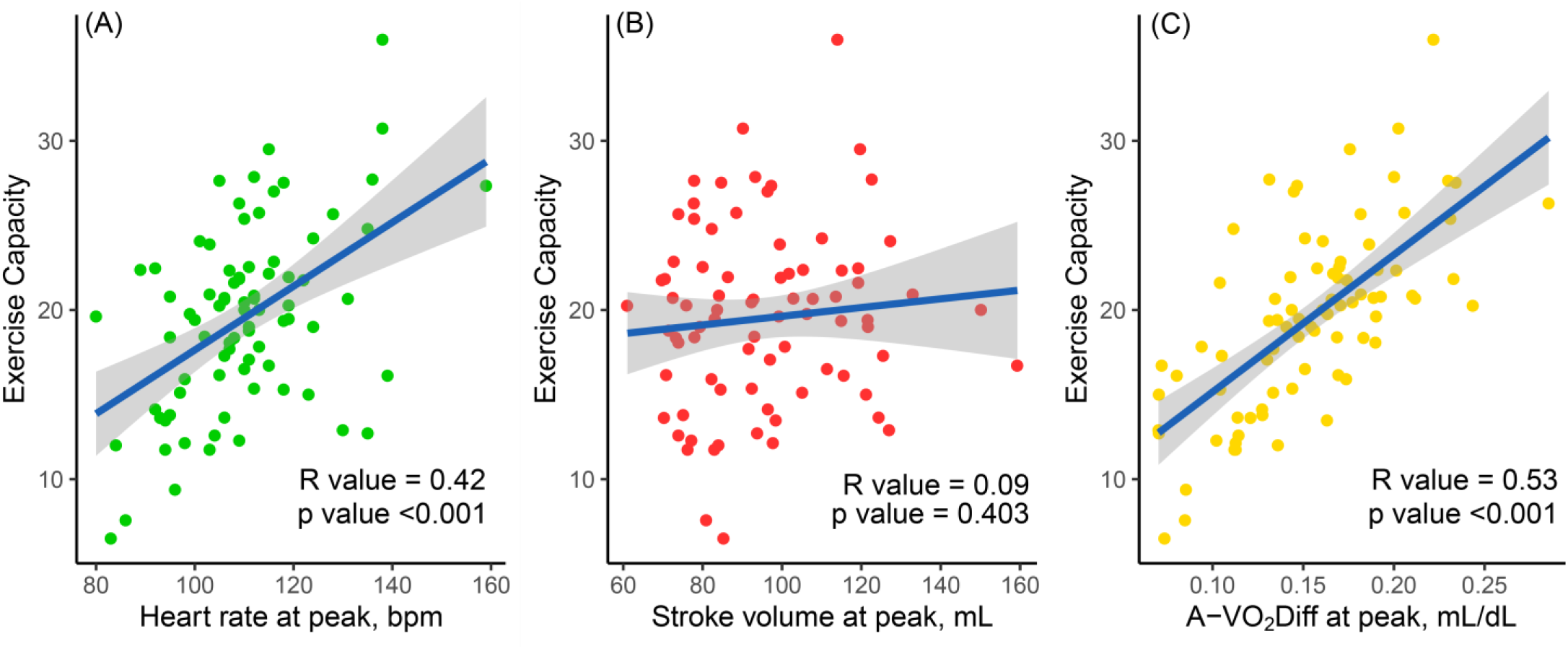
The univariate relationship between exercise capacity (oxygen uptake at peak exercise in mL/min/kg) and variables at peak exercise: heart rate (A), stroke volume (B) and A-VO2Diff (C). The gray-shaded area represents the standard error of regression line (blue). The R value was calculated using Pearson correlation. In contrast to multivariate analysis there is no significant association between stroke volume at peak and EC in the univariate analysis; A-VO2Diff, arteriovenous oxygen difference.

### Exercise capacity after acute myocardial infarction

EC in our studied group is comparable to previous studies of patient entering cardiac rehabilitation. Similarly, to our study in the study by Kavanagh et al, a cohort of post AMI patients entering cardiac rehabilitation tested on the cycle ergometer showed a comparable mean values of peak VO2, 20.5 ± 5.2 mL/kg/min in men and 15.4 ± 4 mL/kg/min in women.1,2 Also in a group of patients after AMI exercised on treadmill before cardiac rehabilitation reported by Ades at al., peak VO2 was 20.4 ± 6.6 mL/kg/min and 14.7 ± 4.2 mL/kg/min in men and women, respectively.^3^ These studies focused on prognostic significance of EC but not on mechanisms leading to low EC.

### Contributors of exercise intolerance

Our findings concur with the results of previous studies on exercise intolerance which described heart rate, stroke volume and skeletal-muscle function as the main contributors to exercise intolerance.^13,14,17,18,33-35^ The contribution of each of these factors to EC varies depending on the individual patient’s disease profile but were not investigated in patients treated for AMI. Results of CPET-SE examination of patients with heart failure have demonstrated exercise intolerance among patients with mid-range and preserved LV EF, to be predominantly influenced by peripheral factors (such as A-VO2Diff), but among patients with reduced LV EF, by decreased stroke volume.^17^

Similarly to our study, peak A-VO2Diff has been found to be the major exercise-limiting factor in patients with heart failure with preserved LV EF evaluated with cardiopulmonary exercise testing with invasive hemodynamic monitoring.^33^

Also, a recently published study involving patients with hypertension with and without heart failure with preserved LV EF found reduced peak VO2 to be related to decreased peak A-VO2Diff. ^18^

In another study of 14 subjects with normal cardiac function, 16 patients with heart failure with preserved LV EF examined for effort intolerance using CPET-SE: heart rate and A-VO2Diff at peak exercise, but not stroke volume, were the most significant independent predictors of EC. However, among patients with heart failure with preserved LV EF, diastolic dysfunction was also found to be a determinant of EC.^14^

Furthermore, similar mechanisms of reduced EC, as in our study, were found in healthy subjects and in patients with HF and preserved or mid-range EF. In a study of 48 patients with heart failure and preserved LV EF assessed with CPET-SE, both reduced cardiac output and A-VO2Diff contributed significantly to exercise intolerance. But, in this study the strongest independent predictor of peak VO2 was the change in A-VO2Diff from rest to peak exercise. ^13^

Although previous studies demonstrated influence of diastolic dysfunction and functional mitral regurgitation on EC,36-38 our study did not confirm these findings. In our studied group only 2 patients have E/e’ ratio at peak exercise >15, which indicates that most of our patient had normal left ventricular end-diastolic pressure during exercise. Also, we did not include patients with severe mitral regurgitation. During exercise, only 7 patients had moderate, and none had severe mitral regurgitation.

### Combined Stress Echocardiography and Cardiopulmonary Exercise Testing

The increased accessibility of CPET-SE provides the opportunity to assess peripheral oxygen extraction during daily clinical practice. CPET-SE is not methodologically standardized. The use of a cycle ergometer in a semi-recumbent position has been suggested to offer improved echocardiographic evaluation.^15,16^ The combination of cardiopulmonary exercise testing with exercise stress echocardiography is a valuable diagnostic tool and its clinical utility has been proven in the diagnostic evaluation of many cardiac diseases including heart failure with reduced, mid-range or preserved LV EF; cardiomyopathies; pulmonary arterial hypertension; valvular heart disease and coronary artery disease.^13,14,17,18,36,39-42^ Furthermore, CPET-SE provides additional information in the case of patients without heart failure, but with unexplained exercise dyspnea.43,44 Exercise pulmonary hypertension due to mitral regurgitation or left ventricular dysfunction can also lead to reduced EC.37,41,42 Furthermore, elevated left ventricular filling pressure during exercise in patients with exercise intolerance and without diastolic dysfunction at rest can be identified by CPET-SE.40 Because resting LV EF is weakly correlated with exercise capacity, there is a need to clarify other parameters contributing to exercise performance including left and right ventricular contractile reserve, interventricular dependence, diastolic function, left atrial function as well as peripheral factors.^37,39,44^

### Limitations

This study has several limitations. We only included patients able to exercise. The mode of exercise used, cycle ergometer in a semi-recumbent position, could cause lower-extremity muscle fatigue, especially in untrained patients. Furthermore, we recruited a relatively small group of patients from a single center. Some patients did not consent to participate in the study introducing selection bias.

### Conclusions

EC improvement after cardiac rehabilitation in patients treated for AMI has been proven in many studies.45-47 In patients with preserved or mildly reduced LV EF it could be mainly due to peripheral factors such as systemic vascular resistance or muscle restoration.^48^

In conclusion, in patients treated for AMI with normal or mildly reduced LV EF, EC is predominantly influenced by peripheral oxygen extraction at peak exercise, but also by peak heart rate and peak stroke volume. Resting left ventricular systolic function and extent of myocardial scarring do not predict EC. These factors can be evaluated using CPET-SE. Our findings can help in clinical decision making in patients after AMI. Strategy based on CPET-SE could potentially be useful in qualification, planning and assessment of the results of cardiac rehabilitation after AMI. The utility of this strategy needs to be validated in further studies.

## Data Availability

The complete raw dataset can be accessed via the Mendeley Data below.

https://data.mendeley.com/datasets/dphcf83ty6/2

## Acknowledgements

This study was supported by the Centre of Postgraduate Medical Education, Warsaw, Poland [grant number: 501-1-10-14-15]. The sponsor did not contribute in study design; in the collection, analysis and interpretation of data; in the writing of the report; and in the decision to submit the article for publication.

## CRediT authorship contribution statement

**Krzysztof Smarz**: Conceptualization, Methodology,

Investigation, Validation, Resources, Data curation, Writing – original draft, Review & editing, Visualization, Project administration, Funding acquisition. **Tomasz Jaxa-Chamiec**: Conceptualization, Writing – review & editing, Supervision, Project administration, Funding acquisition. **Beata Zaborska**: Conceptualization, Methodology, Writing – review & editing. **Maciej Tysarowski**: Conceptualization, Methodology, Software, Formal analysis, Resources, Data curation, Writing – review & editing, Visualization. **Andrzej Budaj**: Conceptualization, Methodology, Writing – review & editing, Supervision, Project administration, Funding acquisition.

All authors take responsibility for all aspects of the reliability and freedom from bias of the data presented and their discussed interpretation. All authors approved of the final version to be submitted.

